# Innate immune responses in patients treated with SBRT irradiation enhances prostate cancer remissions

**DOI:** 10.1101/2022.01.06.22268830

**Authors:** Amrita K Cheema, Yaoxiang Li, Sean Collins, Simeng Suy, Mary Ventimiglia, Keith Kowalczyk, Ryan Hankins, John Lynch, Einsley-Marie Janowski, Scott Grindrod, Alejandro Villagra, Anatoly Dritschilo

## Abstract

Stereotactic body radiation therapy (SBRT) is a curative therapeutic modality employing large fractional doses of highly conformal radiation therapy for cancer treatment. To understand the mechanisms underlying clinical responses to radiation therapy, SBRT offers a unique window for high-throughput analysis of post-radiation molecular events to inform predictive biomarker discovery and strategies for multi-disciplinary therapeutics. We performed longitudinal analysis of plasma proteins and metabolites from patients treated with prostate SBRT, comparing cohorts of patients in clinical remission to cohorts experiencing PSA-determined cancer progression. We observed onset of post-SBRT DNA Damage Response (DDR), cell cycle arrest, and immune response signaling in patients within one hour of treatment and innate immune response signaling that persisted for up to three months following treatment. Furthermore, patients in remission experienced more robust immune responses and metabolite elevations consistent with a pro-inflammatory, M1-mediated innate immune activation in the short-term following SBRT, whereas patients with disease progression had less robust immune responses and M2-mediated metabolite elevations. We interpret these data to support a critical role for innate immune activation in the clinical outcomes of patients receiving radiation therapy for prostate cancer potentially improving future multidisciplinary therapeutic strategies.

**One Sentence Summary:** Following SBRT, proteomic and metabolomic profiles reveal a robust immune activation that correlates with prostate cancer remission

## Introduction

Prostate cancer is a major cause of death and disability in men, with estimates of 248,530 diagnoses and 34,130 deaths in 2021 in the U.S. [1]. Radiation therapy (RT) is an effective modality for curative treatment of prostate cancers as a single agent and in combination with hormonal therapies or after surgery. Efforts to improve outcomes of RT have focused on advances in imaging, beam shaping and dose fractionation. The development of stereotactic body radiation therapy (SBRT) utilizing a robot mounted linear accelerator to deliver precise, highly conformal radiation therapy to the prostate in large fractional doses has yielded excellent clinical outcomes and shortened the overall treatment time from two months to two weeks [2, 3]. Understanding signaling pathways and the molecular mechanisms in patients undergoing SBRT offers the opportunity to gain insight into cellular and systemic responses to RT.

The immune system has been implicated in patients undergoing RT through observations of “abscopal” cancer responses, as well as improved clinical outcomes in recent clinical trials [4-6]. Recent advances in immune directed therapies and personalized medicine have also been extended to treating advanced, metastatic prostate cancers [7]. Observed benefits, risks, and late effects in the heterogeneous clinical responses of patients receiving curative doses of radiation therapy (RT), underscore the complexity of clinical therapeutics and the urgent need to understand the biology. Integrated responses of tumors and normal tissues following radiation therapy enable the discovery of predictive biomarkers and therapeutic molecular targets.

Although cancers confined to the prostate can be cured by radiation therapy, dose limitations of normal tissues at risk and the potential for undiagnosed metastases underlie treatment failures and cancer recurrences. Recent advances in determining roles for the DNA Damage Response (DDR) and cell cycle arrest after radiation exposure have focused on cancer cell sensitization strategies. In addition, reports of abscopal antitumor immune response have implicated a contribution by the immune system to both, local tumor control and regression of metastases. Regardless of the antitumor immune responses generated by RT, irradiated tumors can recruit monocytes to the injured area, which are quickly differentiated into tumor-associated macrophages (TAMs) [8, 9]. TAMs are mainly polarized towards the protumoral M2 phenotype and are strongly associated with a poor prognosis in cancer ([10-13].

TAMs produce anti-inflammatory cytokines, induce hypoxia, express immunosuppressive mediators, and support tumor growth. These functional characteristics of M2 macrophages have a detrimental effect on CD8 effector T cell function [14, 15]. Thus, controlling the recruitment or polarization of macrophages in irradiated tumors could be an attractive option to prevent the activation of survival pathways with RT.

To understand molecular events characterizing the global clinical responses in irradiated patients, we enrolled 132 patients receiving stereotactic body radiation therapy (SBRT) for prostate cancers on an IRB approved protocol to serialy collect blood and quality of life data prior to and after treatment. Plasma protein and metabolite profiles were determined relative to the pre-RT clinical specimens in a time course after SBRT. We then analyzed the global responses using high-throughput proteomics and metabolomics as well as comparisons of cohorts of patients experiencing disease remission to those with cancer progression. We report here the robust immune system signaling in irradiated patients and a strong correlation of innate immune system activation to clinical outcomes.

## Results

SBRT is a radiation modality that utilizes advanced image-based technology for precise targeting and delivery of hypo-fractionated RT over an interval of 1 to 2 weeks [2]. Radiation dose fraction sizes in this study range from 6.5 to 7.25 Gy, doses that are approximately three times greater than conventional daily RT fraction sizes. We previously reported our clinical outcomes of tumor control and radiation late effects using the Accuray CyberKnife, a clinical, robot-mounted linear accelerator system at the MedStar Georgetown University Hospital (M-GUH) [3, 16, 17]. Patients were enrolled into an institutional review board approved, prospective quality of life clinical protocol. We used a multi-omics based molecular phenotyping approach to characterize serum samples from a cohort of patients (N=132) who received SBRT at M-GUH to treat localized prostate cancer. Of the 132 enrolled patients, seventeen patients experienced recurrence as defined by PSA progression [18]. Peripheral blood was drawn before the treatment (Pre), after 1 hour, 24-hour, 1 month, 3 months, 6 months, and 12 months (Figure 1, Panel A). Our patient population included 60% Caucasian and 34% African American males. The clinical data for disease burden assessment, including baseline PSA, biopsy Gleason score, and tumor score were used to assign the subjects to risk categories according to the D’Amico criteria[19]. Briefly, patient mean age was 70 years, mean PSA was 8.6 ng/mL and 28% were high risk, 53% intermediate risk and 16% low risk groups (Figure 1, Panel B). Processed study samples were analyzed by SomaLogic, Inc., using the SOMAscan Version 3 proteomic assay. Relative distribution of significantly dysregulated pathways was evaluated using Doughnut charts, which showed discreet differences within 24 hours of RT (top) with persistent changes at 1-month (bottom) that included early onset of changes in immune response, interleukin signaling, PI3K activated AKT signaling, and MAPK signaling pathways upon RT (Figure 1, Panel C).

**Figure 1.**
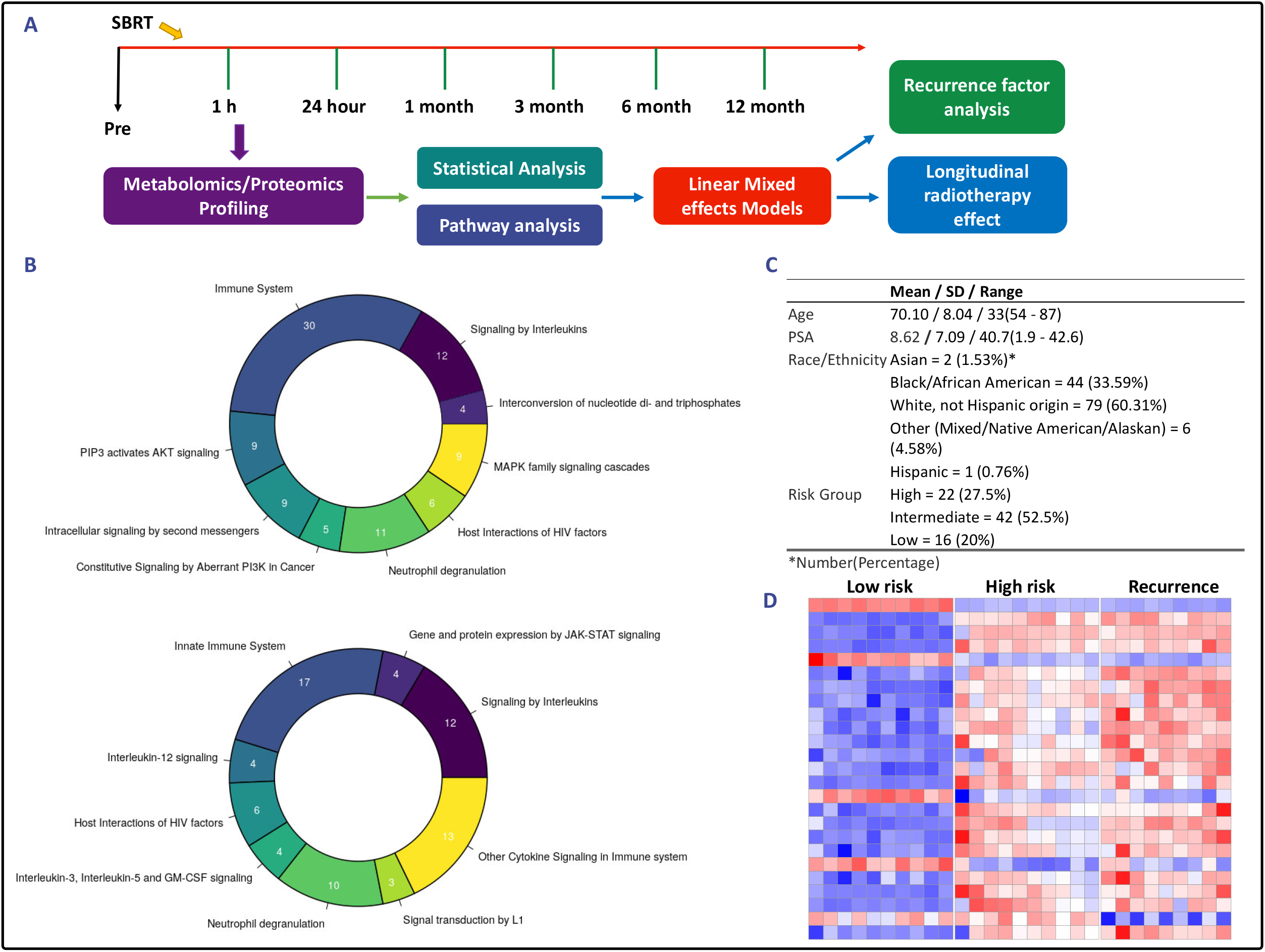
Summary of main findings showing that radiation therapy (RT) induces robust molecular alterations that modulate tumor response in prostate cancer. **Panel A**. Overall study design. Enrolled patients (N=132) were diagnosed with prostate cancer and elected SBRT radiation treatment. Patient plasma samples were obtained before treatment (pre-RT), after 1 hour, 24-hours, 1-, 3-, 6-, and 12-months post-RT. Tumor response to RT was characterized by performing multi-omics analyses (metabolomics, lipidomics and proteomics) of patient plasma samples. **Panel B**. Doughnut chart with proportions of significantly dysregualated proteomics pathway, based on the number of entities found in each pathway showed discreet differences in the 24 hours vs. baseline (top) and 1-month vs. baseline (bottom) groups. **Panel C**. Clinical characteristics of the prostate cancer cohort that received RT for treatment of prostate cancer. Seventeen patients experienced recurrence episodes. **Panel D**. Hierarchical clustering-based heat map visualization of metabolite patterns that segregate prostate cancer patients based on clinical risk group.

Un-paired t-tests were used to determine overall changes in global protein expression at each time point (1 hour, 24 hour, 1 month, 3 month, 6 month, and 12 month) following RT as compared to pre-treatment (baseline) levels (Supplementary Table 1). Reactome pathway analyses [20] were performed for all significantly changed metabolites using their UniProt ID (Supplementary Table 2).

Untargeted metabolomics was performed in a subset of patients classified as low, high and recurrence groups (N= 10 each) and further validated through tandem mass spectrometry (MS/MS) for selected metabolites (Supplementary Table 3). Un-paired t-tests were conducted to study overall radiation response (Supplementary Table 4) while linear mixed effects models were used to identify molecular determinants of tumor response using a retrospective clinical outcome analysis (Supplementary Table 5). Hierarchical clustering-based heat map visualization showed distinctive patterns of metabolic abundance in plasma amongst low, high and recurrence risk groups as scored by current clinical criteria, suggesting distinct metabotypes that were worthy of further investigations (Figure 1, Panel D).

### DDR and innate immune response could be an early indicator of tumor outcomes of RT

We used Reactome analysis to interrogate the longitudinal proteomics data set to gain insights into pathway perturbations following RT. Interestingly, we found that DDR, cell cycle arrest, and immune response signaling activated within one hour after RT, DDR and cell cycle activation were short-lived and waned by the 1-month post-RT time point while immune activation persisted for up to 3 months (Figure 2, Panels A and B). These observations suggest that robustness of the immune signaling response in this analysis was greater than that of either DDR or cell cycle arrest. Next, we asked if immune response activation correlated with PSA determined tumor recurrence; hence, we used linear mixed effects models to identify significantly dysregulated proteins. Herein, we use “time” as a random effect and “recurrence” as a fixed effect to determine significant differences between recurrence groups adjusted for time for each protein as an outcome measurement (Supplementary Table 6). Examination of patterns of serum protein abundance revealed increased expression of DDR and immune response proteins including interferon gamma, proteasome subunit alpha and ubiquitin-conjugating enzyme among others within an hour of RT in patients that went into remission while the serum abundance remained relatively unchanged (as compared to baseline) in patients with clinical recurrence (Figure 2, Panel C). Western blot analysis of CD86 showed an initial increase in serum levels in the remission group as compared to the recurrence group that showed minimal change suggesting that an onset of a M1 macrophage (inflammatory) phenotype following RT could be an early determinant of tumor response (Supplementary Figures 1A and 1B). Since radiation damage to DNA and the tumor microenvironment (TME) underlie molecular and cellular processes that induce DDR, arrest cell cycle progression, and activate the immune system, these results suggest that DDR and Immune response may likely modulate tumor response to RT.

**Figure 2.**
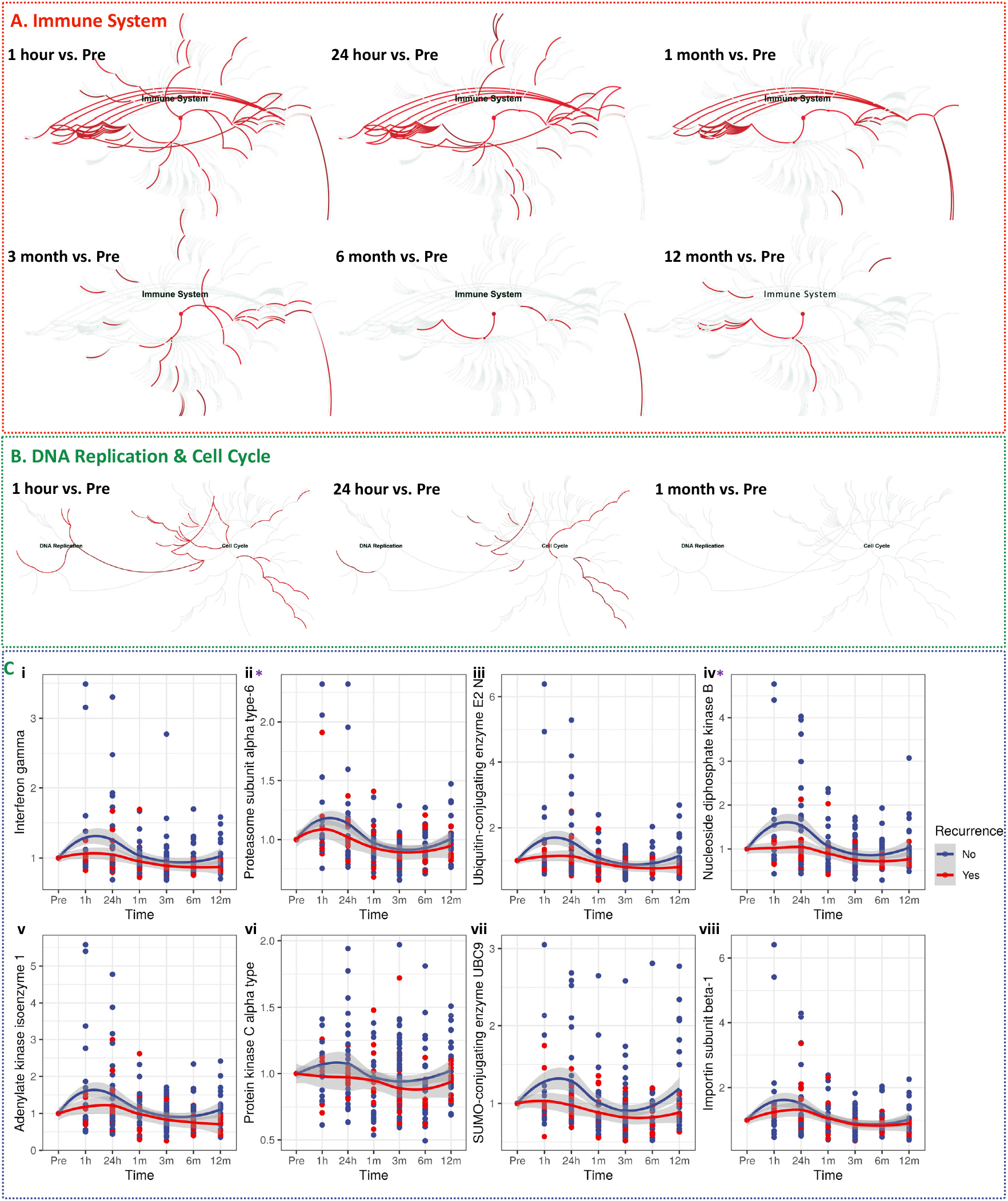
Radiation induced early immune response is associated with tumor response in prostate cancer. **Panel A**. Reactome based longitudinal pathway analysis of the plasma proteomics data set. A robust immune signaling response was observed within an hour of SBRT and the duration of signaling extended longer than DDR or cell cycle signaling. Analysis of selected markers of immune response shows a more muted innate immune response in patients with cancer recurrence. **Panel B**. Reactome based pathway analysis shows activation of DDR and cell cycle within 1 hour after RT, attenuation of the response by 24 hours and return to pre-RT baseline by 1 month. **Panel C**. Trend lines showing differential pattern of protein expression changes over time for immune response (sub-panels i - iv), DDR and cell cycle (sub-panels iv - viii), in patients undergoing remission (blue) as compares to the recurrence group (red). Proteins overlapping across different functional groups are marked with asterix*.

### Anti-inflammatory metabotype correlates with biochemical recurrence in prostate cancer

To validate the proteomics data, we analyzed the metabolomic profiles to determine the segregation of key molecules associated with macrophage metabolism. Figure 3, panel A shows principal component analysis (PCA)-based separation of patient cohorts by risk of recurrence in pre-SBRT and post-SBRT clinical samples. Macrophages can be classified according to their inflammatory phenotype into proinflammatory M1 and anti-inflammatory M2, which are known to promote an immunosuppressive or immunoreactive TME, respectively [21-23]. Macrophage phenotype in the TME correlates with the aggressiveness in most types of cancer [24, 25]. M1 and M2 macrophages exhibit distinct metabolic types; for example, M1 macrophages are glycolytic and break down amino acid arginine to nitric oxide, while the M2 macrophages produce ornithine and uric acid [26]. We found that expression markers and metabolic phenotype of the M2 macrophages are upregulated in prostate cancer patients with progressive disease after RT (Figure 3, Panel B and Supplementary Table 5) thus confirming the correlation of immunosuppressive response associated metabolites in predicting prostate cancer recurrence. Plasma levels of metabolites including citric acid, ornithine and uric acid (produced by M2 macrophages) were elevated in high-risk and recurrence groups post-RT as compared to the low-risk groups although baseline levels of these metabolites were comparable in all three groups (Figure 3, Panel C). This suggests that tumor failure following RT, at least in part, could be attributed to an anti-inflammatory immune-metabolic phenotype.

**Figure 3.**
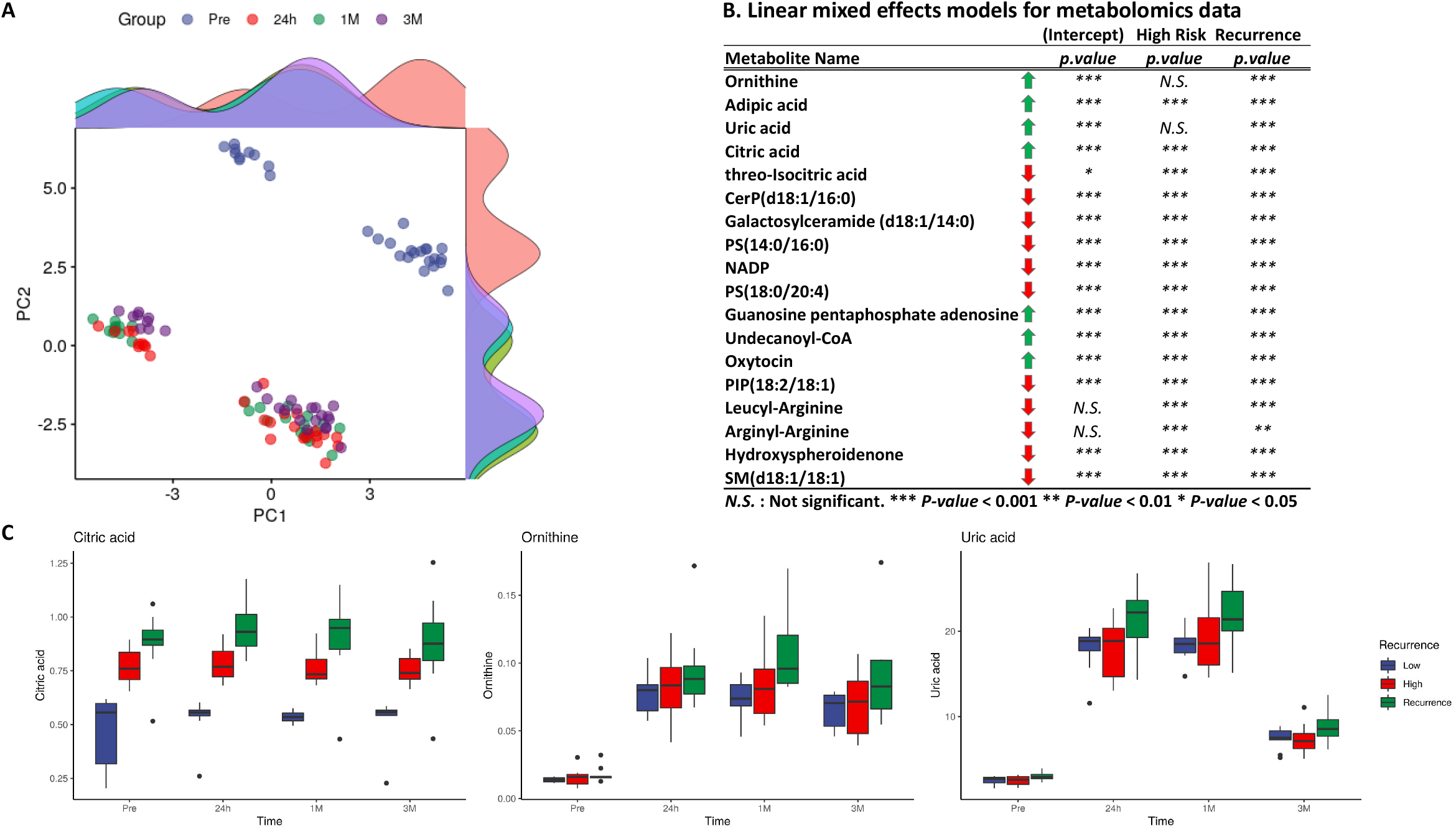
Metabolomic profiles can segregate PC patients based on recurrence. **Panel A**. 2D-PCA for metabolomics data showing separation for Pre, 24-hour, 1 month, and 3-month samples. **Panel B**. Linear mixed effects models for longitudinal metabolomics using “time” as a random effect and tumor recurrence as a fixed effect, to see are there any significant differences between Recurrence groups adjusted for time for each protein as outcome measurement. Metabolites associated with immune response are significantly dysregulated. **Panel C**. Box plots showing metabolites associated with M2 macrophage phenotype are upregulated in patients that experienced recurrence.

## Discussion

A significant percentage of cancer patients (50-55%) receive radiation therapy (RT) either alone or in combination with surgery or chemotherapy during the course of their treatment. However, the biological and molecular basis of tumor response to radiation remain understudied areas of research. Ionizing radiation induces various molecular, cellular, and biological effects both directly by interacting with DNA or indirectly by forming free radical species that damage DNA. In normal and transformed cells, the resultant biochemical and molecular signals may lead to the expression of specific DDR genes, protein modifications, activation of metabolic reactions, generation of pattern recognition receptors (PRRs), and induction of cell surface antigens. In turn, the signaling cascades may result in activation of cellular pathways (apoptosis, necrosis or autophagy) and the innate immune system to reshape the composition of the TME [27-29].

Radiation therapy of prostate cancers offers a window into the local regional effects on tumors as well as the systemic immune pathway activation. A better understanding of the role of the immune response informs predictive biomarkers and identifies therapeutic targets to enhance the effort to cure these cancers. In this clinical investigation we applied state-of-the art molecular analyses and big data analysis to gain insight into cancer and host responses to SBRT.

We used metabolomics and proteomics-based molecular phenotyping approach to study tumor response to radiation in a cohort of 132 prostate cancer patients treated with Stereotactic Body Radiation Therapy (SBRT) at Medstar-Georgetown University Hospital (M-GUH) in a longitudinal, quality-of-life study. We discovered that activation of DNA damage response (DDR) pathways upon irradiation of tumor and the subsequent activation of innate immune response strongly correlated with tumor regression in our cohort of prostate cancer patients, a finding that corroborates studies reported by others with murine models [30, 31].

Plasma metabolomics helped corroborate these findings suggesting immunometabolic activation may play a critical role in dictating tumoral response to RT. Several recent publications have reported that the presence of M2 macrophages is directly associated with poor prognosis in cancer, through enhancement of tumor immune-evading mechanisms [24, 25, 32-34]. Additionally, this association can be extended to patients treated with immunotherapy and targeted therapies [35-37]. However, the correlation between the M1/M2 macrophage ratio and improved prognosis in cancer has not been described comprehensively in the context of RT.

Recent findings have identified a critical role of the TME cellular composition after exposure to ionizing radiation. In this context, RT initially triggers activation of proinflammatory, anti-tumor M1 macrophages, followed by the recruitment of tumor-associated macrophages (TAMs) that predominantly exhibit the M2 phenotype [38-40]. Additionally, macrophages are critical modulators of the metabolic landscape in tumors, a key component of cancer aggressiveness [41, 42]. Therefore, the ratio of antitumoral M1 and protumoral M2 macrophages (M1/M2) have been proposed as a potential biomarker for various malignancies, including prostate cancer [25, 43-45]. Thus, our work offers a new perspective and possibilities to identify more accurate and significant markers of responses pre- and post-SBRT. Variations in clinical sensitivities to radiation have been observed in patients with genetic syndromes, mutations in genes associated with DNA repair processes, cell cycle checkpoints and immunological diseases [46].

In summary, RT induces DNA damage, activating cellular processes that lead to DNA damage response (DDR) mechanisms and DNA repair. Cellular injuries in the irradiated tumor microenvironment (TME) also trigger activation of the innate immune system and recruitment of an orchestrated pool of immune cells. As such, systematic studies of molecular interactions and cellular crosstalk underlying immune responses, DDR, and TME activation by RT are imperative to understand the biologic basis of radiation response to improve therapeutic strategies in RT-based cancer treatment. This would help advance personalized medicine in radiation oncology, optimizing radiation treatment planning based on the individual patient’s unique biological signatures.

## Materials and Methods

### Patient Recruitment and Study population

Patients were enrolled at MedStar–Georgetown University Hospital (GUH) into IRB protocol 2012-1175; an approved quality of life clinical trial. The protocol permits longitudinal collection of clinical samples, symptom monitoring and quality of life data which have contributed to interim published reports of clinical outcomes including GU and GI acute and late effects [2, 7, 17]. This study population is a part of ongoing recruitment of PC patients coming in through the referral network to GUH. The study participants include men of Caucasian, African American, Asian and Hispanic descent, as well as men of other ethnicities, aged 35-70 years, residing in Washington DC and surrounding areas, who were diagnosed with localized prostate cancer by biopsy. Patients are recruited from the Departments of Radiation Medicine and Urology at the GUH. Approximately 150 newly diagnosed patients with prostate cancer receive SBRT for prostate cancer each year. All protocol enrolled participants complete informed consent for blood and urine collection and periodic self-reported symptom monitoring. Prior to enrollment, all patients undergo physical examination, including a digital rectal examination (DRE). Phlebotomy (by trained phlebotomists) and urine collection are performed before treatment and at each following visit thereafter (1, 3, 6, 9 and 18 months) after SBRT treatment. Patient eligibility criteria include histologically confirmed adenocarcinoma of the prostate (biopsy within 1 year of enrollment); Gleason score 2-10; clinical stages T1c-T3c; no clinically or pathologically involved lymph nodes on imaging; no distant metastases on bone scan; measurement of prostate serum antigen (PSA) levels <60 days prior to registration; no history of pelvic radiotherapy, chemotherapy, or radical prostate surgery; no recent (within 5 years) or concurrent cancers other than non-melanoma skin cancers; no medical or psychiatric illnesses that would interfere with treatment or follow-up; a baseline AUA/IPSS score of < 20; and no history of inflammatory bowel disease. All patients sign a study-specific consent form. Patients complete a detailed questionnaire regarding familial cancer history, tobacco use, medication use, occupational history and socioeconomic status, the 26-item EPIC (sexual, bowel, and urinary symptoms). Other patient data such as patient de-identifier number, prostatic volume, Gleason’s grade, prior hormonal therapy, clinical co-morbidities, age, ethnicity, body mass index etc. were recorded. Blood samples are processed for serum and plasma collection. In this study we have used plasma samples for metabolomic profiling. Buffy coat, mononuclear cells and RBCs are also collected and stored for future studies. All samples are processed within four hours of collection, aliquoted and stored at -80°C within 4 h of collection to preserve sample integrity.

### Proteomic Analysis

Serum samples were analyzed on the proteomic discovery platform described by Gold et al [47]. Briefly, this technology uses novel DNA aptamers, which are chemically modified nucleotides, to act as highly specific protein binding reagents, thereby transforming the quantify of each targeted protein into a custom hybridization array. Protein quantities were recorded as relative fluorescent units (RFU), which can be converted to concentrations with standard curves. The samples were batch processed using the SOMAscan Version 3 assays according to the manufacturer’s instructions. This assay is commercially available and has been used to investigate other disease systems including lung cancer. In this study, 1129 protein targets were measured in 15 µL of serum for each subject, and all sera were analyzed in a continuous process. All samples were normalized and calibrated using standard procedures prior to analysis. The identity of the samples was completely blinded throughout the proteomic analysis process. Pathway analysis was performed using Reactome [20]. After data pre-processing and normalization, the proteomics data was log transformed. Statistical analysis was performed by unpaired t-test using R (Version 4.0.2).

### Mass Spectrometry Based Metabolomic Profiling

Metabolite extraction was performed using 25 µL of plasma sample which was mixed with 175 µL of 40% acetonitrile in 25% methanol and 35% water containing internal standards (10 µL of debrisoquine (1mg/mL), 50 µL of 4, nitro-benzoic acid (1mg/mL), 27.3ul of Ceramide (1mg/ml) and 2.5ul of LPA (4mg/ml) in 10 mL). The samples were incubated on ice for 10 minutes and centrifuged at 14,000 rpm at 4°C for 20 minutes. The supernatant was transferred to a fresh tube and dried under vacuum. The dried samples were resuspended in 200 µL of 5% methanol, 1% acetonitrile, 94% Water. Samples were centrifuged again at 13,000 rpm for 20 minutes at 4°C and the supernatant was transferred to MS vials for UPLC-ESI-Q-TOF-MS analysis. Each plasma sample (2 μL) was injected onto a reverse-phase CSH C18 1.7µM 2.1×100mm column using an Acquity G2-QTOF system (Waters Corporation, USA). The gradient mobile phase comprised of water containing 0.1% formic acid solution (A), 100% acetonitrile (B) and 10% acetonitrile in IPA containing 0.1% formic acid and 10mm Ammonium format (D). Each sample was resolved for 13 min at a flow rate of 0.5 ml/min for 8 min and then 0.4 ml/min at 8 to 13 min. The G2-QTOF gradient consisted of 98% A and 2% B for 0.5 min then a ramp of curve 6 to 60% B and 40% A from 0.5 min to 4.0 min, then a ramp of curve 6 to 98% B and 2% A from 4.0 to 8.0 min, then a ramp of curve 6 to 5% B and 95% D from 9.0 min to 10.0 min at 0.4ml/min, followed by 98% A and 2% B from 11.0 min to 13 minutes. The column eluent was introduced directly into the mass spectrometer by electrospray. Mass spectrometry was performed on a performed on a Q-TOF instrument (Xevo G2 QTOF, Waters Corp, USA) operating in either negative (ESI-) or positive (ESI+) electrospray ionization mode with a capillary voltage of 3200 V in positive mode and 2800 V in negative mode, and a sampling cone voltage of 30 V in both positive and negative modes. Pooled quality controls were analyzed throughout the batch to assess chromatographic reproducibility and data consistency.

### Mass Spectrometry Based Metabolomic Analysis

The untargeted metabolomics data was initially normalized by internal standard. Following data pre-processing and ion annotation, the m/z values of the measured metabolites are normalized with log transformation that stabilizes variance. Differential expression between various patient groups is assessed using t-test constrained by p-value <0.05. Among these differentially expressed metabolites, each m/z value is scored for annotation against the HMDB, Metlin, MMCD and Lipid Maps databases within a 5ppm mass tolerance. We adjust for multiple comparisons using the Bonferroni correction. The heat maps were generated for the significant metabolites using the log2 transformed values of fold changes and hierarchically clustered by Pearson correlation. Statistically significant metabolites were validated using tandem mass spectrometry-based fragmentation matching.

### Western blot analysis

Patient plasma samples were immune-depleted of fourteen high-abundance proteins in human plasma using the Multiple Affinity Removal Column Human 14 (Agilent, USA). Samples were then rebuffered in MPER lysis buffer. BCA protein estimation assay was performed in all the samples to quantify protein. LDS sample buffer, DTT, and DNase/RNase-free distilled water were added to the lysates (30µg of protein in 30µL total volume) prior to denaturing (samples were boiled for 10 minutes) before being separated by electrophoresis using 4-12% NuPAGE Bis-Tris gel (cat. NP0322BOX; Invitrogen). Western blotting for human sera was performed using methods previously described [48, 49]. Briefly, equal amounts of serum protein (30ug; Supplementary Figure 1) were loaded and resolved in each lane, transferred to a PVDF membrane (iBlot 2 Transfer Stacks, cat. IB24002; Invitrogen) and washed with Ponceau S to confirm the successful transfer of proteins to the membranes (Supplementary Figure 1B). The membranes were then washed with TBST and blocked for 1 hour (5% milk in TBST). Next, the membranes were incubated overnight at 4°C with a CD86 polyclonal antibody (dilution 1:500; cat. 13395-1-AP, ProteinTech). Primary antibodies were detected using horseradish peroxidase-conjugated secondary antibodies to rabbit IgG (1:5000). Membrane images were captured using Amersham Imager 600 and band intensity was determined through densitometric analysis using the Image J program.

## Data Availability

All data produced in the present study are available upon reasonable request to the authors

## Author Contributions

Conceptualization: AKC, AD; Methodology: SC, SG, EJ, SS, MV, AKC, AD; Investigation: SG, AV, KK, RH, JL, AKC, AD; Visualization: YL; Funding acquisition: AKC, AD; Project administration: AKC, AD; Supervision: AKC, AD; Writing – original draft: AKC, AD, YL; AV; Writing – review & editing: All authors contributed to review and edits of the manuscript

## Acknowledgements

The authors would like to acknowledge the Metabolomics Shared Resource at Georgetown University Medical Center, partially supported by NIH/NCI/CCSG grant P30-CA051008. We also acknowledge support by Somalogic Inc. for generating Proteomics data vide a collaborative data transfer and research agreement.

## Funding

The study was supported by NIH SBIR (Phases I and II) contract # HHSN261201600027C to Shuttle Pharmaceuticals, Inc.

## Competing interests

Anatoly Dritschilo, MD, owns equity and serves as CEO of Shuttle Pharmaceuticals, Inc. Sean Collins, MD, PhD, receives grant support from Accuray, Inc. Accuray Inc. provided a grant to support a data manager.

## Figure Legends

**Supplementary Figure 1A.**
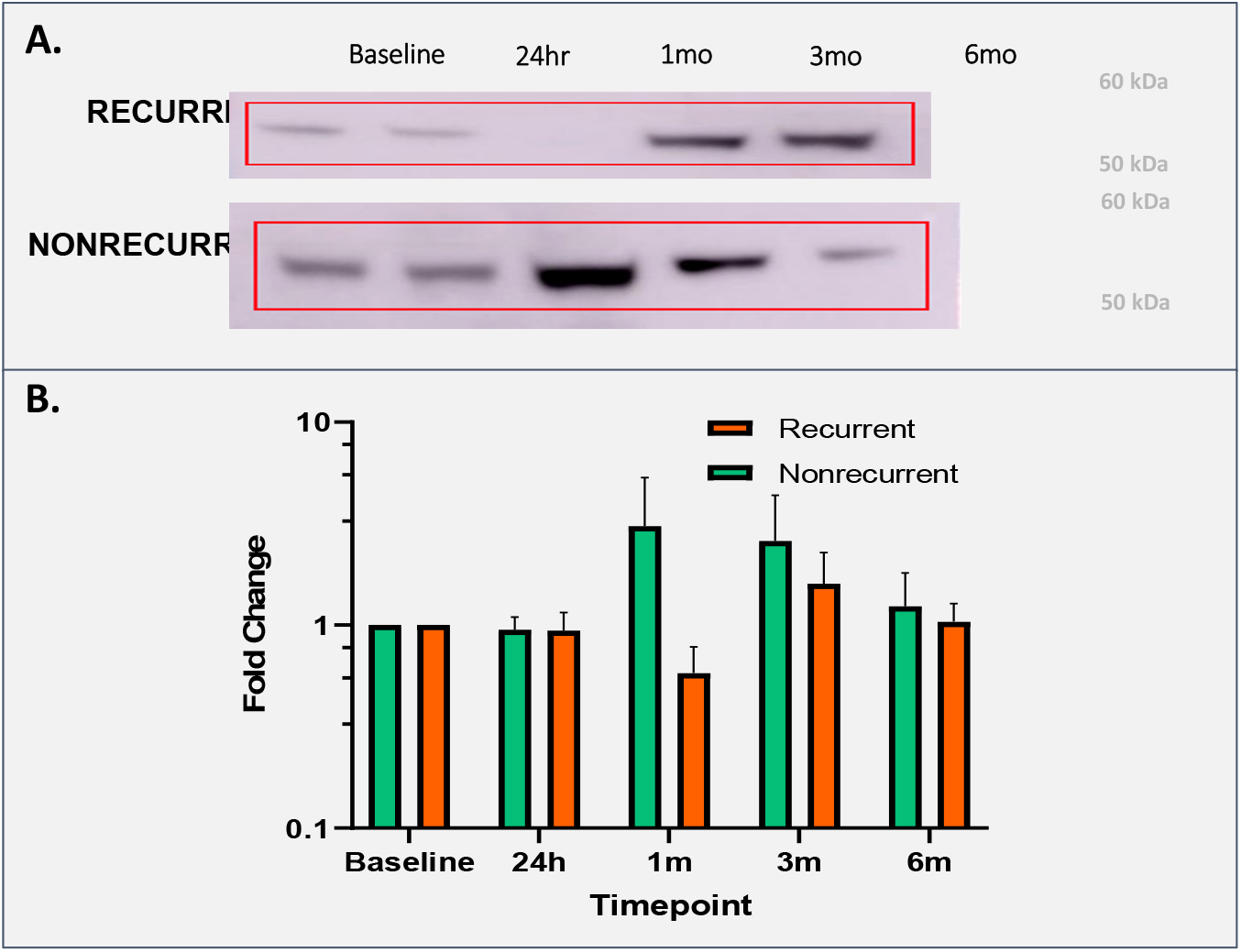
Western blot analysis shows upregulation of M1 macrophage marker CD86 in prostate cancer patients showing remission after RT. (**Panel A**). Albumin-depleted serum lysate samples for 10 prostate cancer patients (five each from the recurrence and non-recurrence groups) were analyzed via SDS-PAGE and western blotting with M1 macrophage polarization marker CD86. Representative (n=10) western blot depicting anti-CD86 hybridization from sera of two post-radiation prostate cancer patients with cancer recurrence and non-recurrence at baseline, 24 hours, 1 month, 3 months and 6 months. (**Panel B**). Average CD86 expression fold-change values from baseline to respective timepoints in recurrence (n=5) cohort and non-recurrence cohort (n=5). At 1-month post-radiation, average CD86 expression in the non-recurrence cohort was ∼3.1-fold greater than baseline whereas expression was 0.6-fold baseline patient expression.

**Supplementary Figure 1B.**
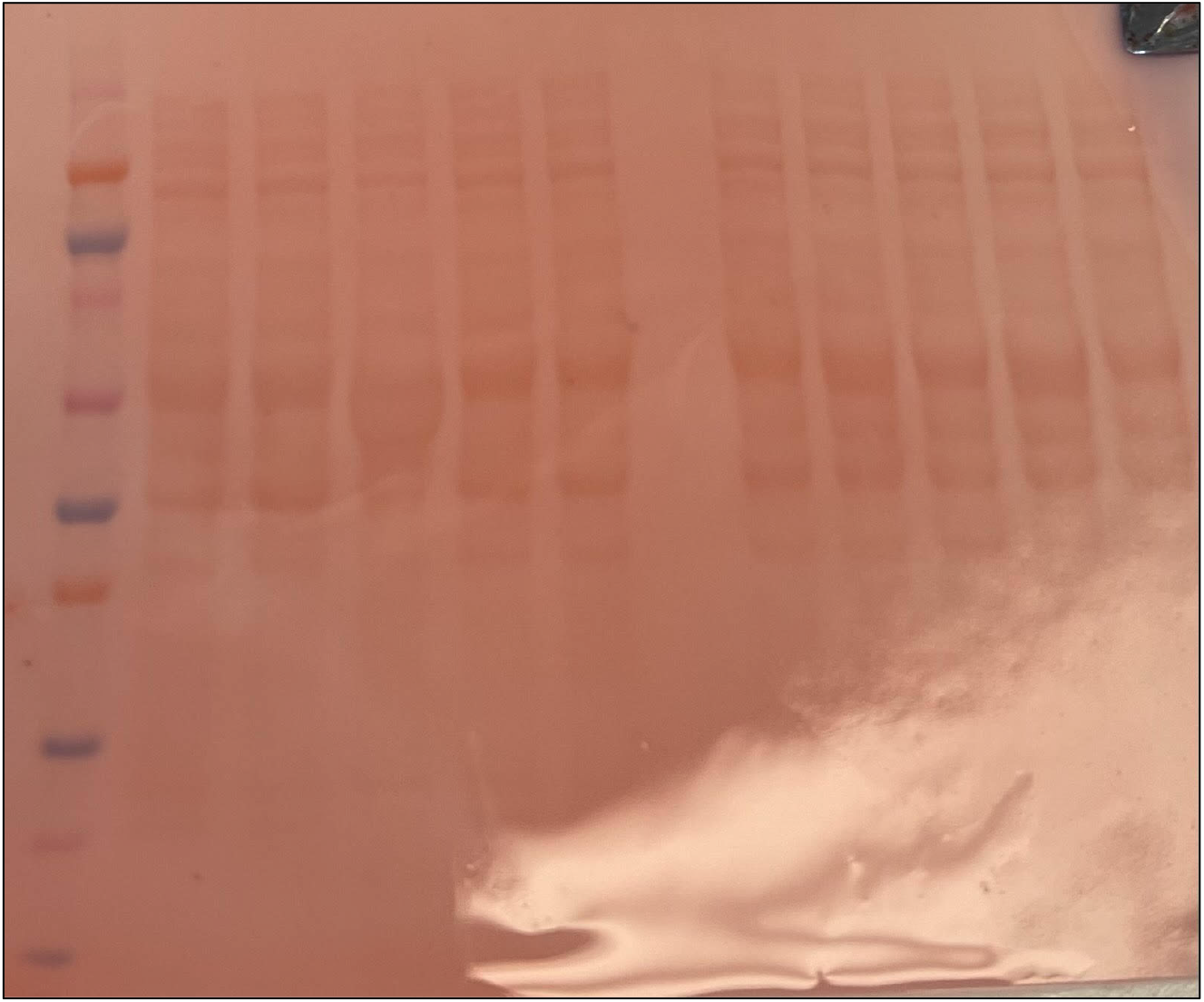
Ponceau S staining to visualize protein loading and successful membrane transfer.

**Supplementary Figure 2.**
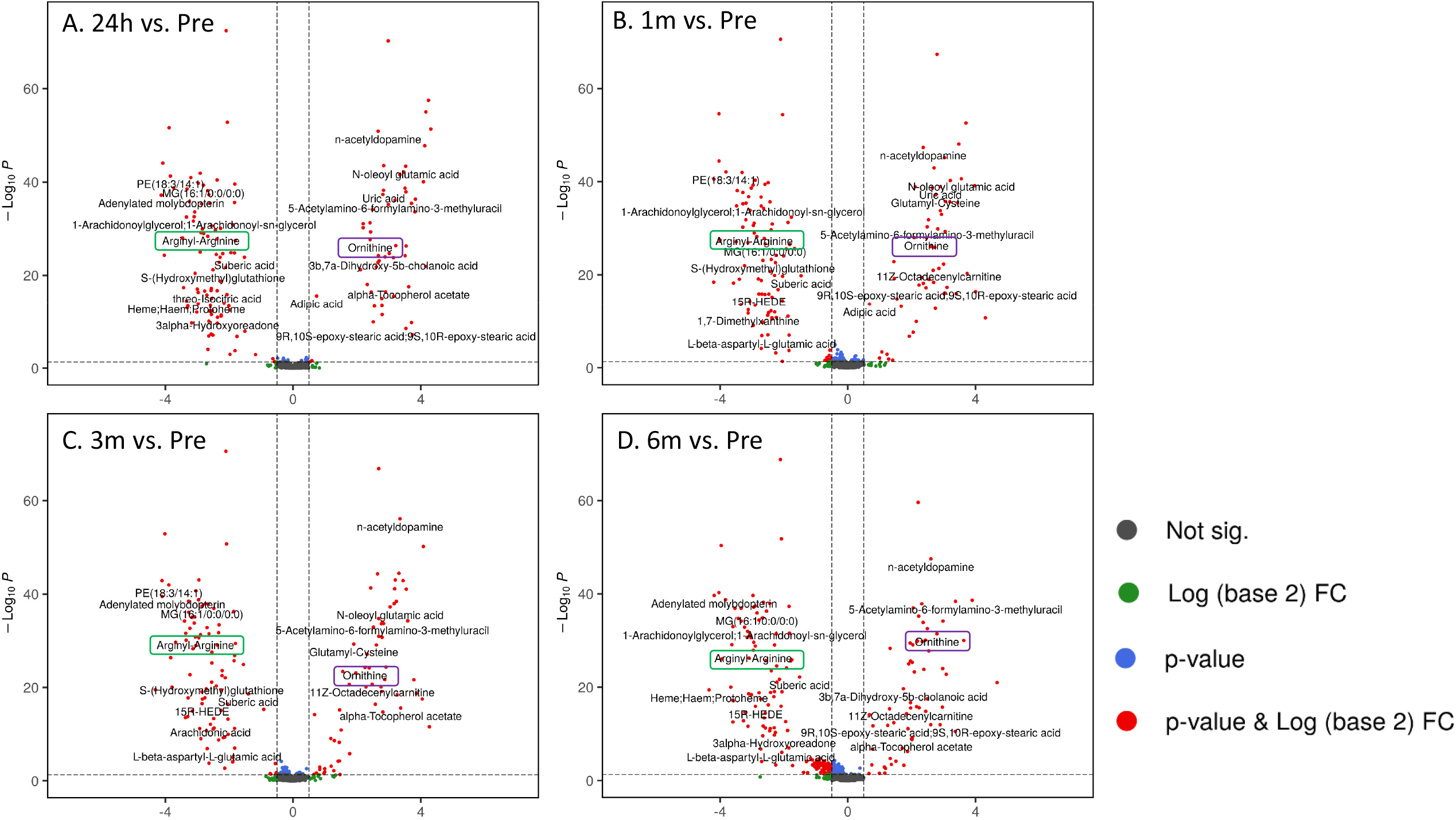
Volcano plot showing significant dysregulation of plasma metabolites at 24 hours (Panel A), one month (Panel B), 3 months (Panel C) and six months (Panel D) as compared to baseline (pre-RT). The red dots depict metabolites with a significant p-value (< 0.05) and fold change (< 0.5 or > 2).

**Supplementary Figure 3.**
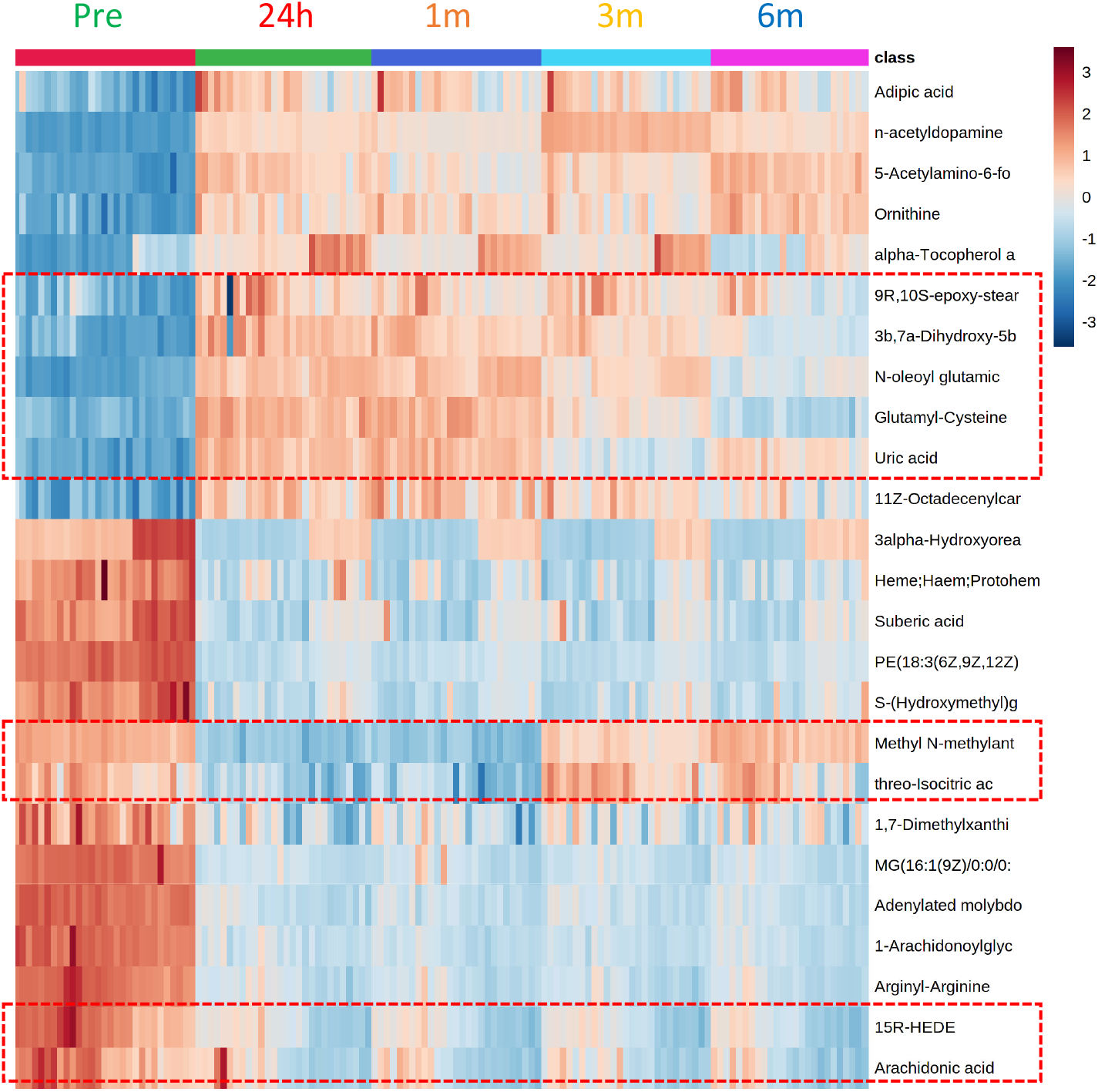
Heatmap visualization of significantly dysregulated following RT. The metabolites were validated using fragmentation matching of tandem MS spectra.

**Supplementary Table 1**. Overall longitudinal (24 hours to 24 months) test statistics for proteomics data set comparing each time post-RT to baseline for the prostate cancer cohort.

**Supplementary Table 2**. Reactome based longitudinal pathway analysis showing significantly dysregulated pathways at 1 h, 24 hours, 1-, 3- and 6-months following RT in the GU prostate cancer cohort.

**Supplementary Table 3**. Validation of metabolite identification using tandem mass spectrometry-based fragmentation matching.

**Supplementary Table 4**. Test statistics comparing each post-RT time point to baseline (pre) in the GU cohort.

**Supplementary Table 5**. Linear mixed effects model analysis of metabolomics data augments validation of immune response findings with proteomics analyses.

**Supplementary Table 6**. Linear Mixed Effect Models analysis of proteomics data combining all time points post-RT.

## Notes

### Competing Interest Statement

The authors have declared no competing interest.

### Funding Statement

The study was funded by NCI/NIH SBIR (Phases I and II) contract # HHSN261201600027C to Shuttle Pharmaceuticals, Inc.

### Author Declarations

Ethics committee/IRB of MedStar-Georgetown University Hospital gave ethical approval for this work

## References

1. American Cancer Society. Cancer Facts & Figures 2021. Atlanta: American Cancer Society.

2. Chen, L.N., et al., Stereotactic body radiation therapy (SBRT) for clinically localized prostate cancer: the Georgetown University experience. Radiat Oncol, 2013. 8: p. 58.

3. Kishan, A.U., et al., Long-term Outcomes of Stereotactic Body Radiotherapy for Low-Risk and Intermediate-Risk Prostate Cancer. JAMA Netw Open, 2019. 2(2): p. e188006.

4. Formenti, S.C. and S. Demaria, Systemic effects of local radiotherapy. Lancet Oncol, 2009. 10(7): p. 718–26.

5. Ngwa, W., et al., Using immunotherapy to boost the abscopal effect. Nat Rev Cancer, 2018. 18(5): p. 313–322.

6. Reynders, K., et al., The abscopal effect of local radiotherapy: using immunotherapy to make a rare event clinically relevant. Cancer Treat Rev, 2015. 41(6): p. 503–10.

7. Cha, H.R., J.H. Lee, and S. Ponnazhagan, Revisiting Immunotherapy: A Focus on Prostate Cancer. Cancer Res, 2020. 80(8): p. 1615–1623.

8. Shiao, S.L., et al., TH2-Polarized CD4(+) T Cells and Macrophages Limit Efficacy of Radiotherapy. Cancer Immunol Res, 2015. 3(5): p. 518–25.

9. Pathria, P., T.L. Louis, and J.A. Varner, Targeting Tumor-Associated Macrophages in Cancer. Trends Immunol, 2019. 40(4): p. 310–327.

10. Zhang, Q.W., et al., Prognostic significance of tumor-associated macrophages in solid tumor: a meta-analysis of the literature. PLoS One, 2012. 7(12): p. e50946.

11. Datta, M., et al., Reprogramming the Tumor Microenvironment to Improve Immunotherapy: Emerging Strategies and Combination Therapies. Am Soc Clin Oncol Educ Book, 2019. 39(39): p. 165–174.

12. Kowal, J., M. Kornete, and J.A. Joyce, Re-education of macrophages as a therapeutic strategy in cancer. Immunotherapy, 2019. 11(8): p. 677–689.

13. Fridman, W.H., et al., The immune contexture in cancer prognosis and treatment. Nat Rev Clin Oncol, 2017. 14(12): p. 717–734.

14. Yu, J., et al., Myeloid-derived suppressor cells suppress antitumor immune responses through IDO expression and correlate with lymph node metastasis in patients with breast cancer. J Immunol, 2013. 190(7): p. 3783–97.

15. Qiu, S.Q., et al., Tumor-associated macrophages in breast cancer: Innocent bystander or important player? Cancer Treat Rev, 2018. 70: p. 178–189.

16. Joh, D.Y., et al., Proctitis following stereotactic body radiation therapy for prostate cancer. Radiat Oncol, 2014. 9: p. 277.

17. Repka, M.C., et al., Predictors of acute urinary symptom flare following stereotactic body radiation therapy (SBRT) in the definitive treatment of localized prostate cancer. Acta Oncol, 2017. 56(8): p. 1136–1138.

18. Paller, C.J. and E.S. Antonarakis, Management of biochemically recurrent prostate cancer after local therapy: evolving standards of care and new directions. Clinical advances in hematology & oncology : H&O, 2013. 11(1): p. 14–23.

19. D’Amico, A.V., et al., Biochemical outcome after radical prostatectomy, external beam radiation therapy, or interstitial radiation therapy for clinically localized prostate cancer. JAMA, 1998. 280(11): p. 969–74.

20. Fabregat, A., et al., Reactome pathway analysis: a high-performance in-memory approach. BMC Bioinformatics, 2017. 18(1): p. 142.

21. Herbst, R.S., et al., Predictive correlates of response to the anti-PD-L1 antibody MPDL3280A in cancer patients. Nature, 2014. 515(7528): p. 563–7.

22. Mantovani, A., et al., Tumour-associated macrophages as treatment targets in oncology. Nat Rev Clin Oncol, 2017. 14(7): p. 399–416.

23. Knox, T., et al., Selective HDAC6 inhibitors improve anti-PD-1 immune checkpoint blockade therapy by decreasing the anti-inflammatory phenotype of macrophages and down-regulation of immunosuppressive proteins in tumor cells. Sci Rep, 2019. 9(1): p. 6136.

24. Zhang, J., et al., Meta-Analysis of the Prognostic and Clinical Value of Tumor-Associated Macrophages in Hepatocellular Carcinoma. J Invest Surg, 2021. 34(3): p. 297–306.

25. Cassetta, L., et al., Human Tumor-Associated Macrophage and Monocyte Transcriptional Landscapes Reveal Cancer-Specific Reprogramming, Biomarkers, and Therapeutic Targets. Cancer Cell, 2019. 35(4): p. 588–602 e10.

26. Viola, A., et al., The Metabolic Signature of Macrophage Responses. Frontiers in Immunology, 2019. 10(1462).

27. Nastasi, C., L. Mannarino, and M. D’Incalci, DNA Damage Response and Immune Defense. Int J Mol Sci, 2020. 21(20).

28. Constanzo, J., et al., Radiation-Induced Immunity and Toxicities: The Versatility of the cGAS-STING Pathway. Front Immunol, 2021. 12: p. 680503.

29. Taffoni, C., et al., Nucleic Acid Immunity and DNA Damage Response: New Friends and Old Foes. Front Immunol, 2021. 12: p. 660560.

30. Markovsky, E., et al., An Antitumor Immune Response Is Evoked by Partial-Volume Single-Dose Radiation in 2 Murine Models. Int J Radiat Oncol Biol Phys, 2019. 103(3): p. 697–708.

31. Yoshimoto, Y., et al., Radiotherapy-induced anti-tumor immunity contributes to the therapeutic efficacy of irradiation and can be augmented by CTLA-4 blockade in a mouse model. PLoS One, 2014. 9(3): p. e92572.

32. Raiha, M.R. and P.A. Puolakkainen, Tumor-associated macrophages (TAMs) as biomarkers for gastric cancer: A review. Chronic Dis Transl Med, 2018. 4(3): p. 156–163.

33. Jeong, H., et al., Tumor-Associated Macrophages as Potential Prognostic Biomarkers of Invasive Breast Cancer. J Breast Cancer, 2019. 22(1): p. 38–51.

34. Kouketsu, A., et al., Regulatory T cells and M2-polarized tumour-associated macrophages are associated with the oncogenesis and progression of oral squamous cell carcinoma. Int J Oral Maxillofac Surg, 2019. 48(10): p. 1279–1288.

35. Tong, N., et al., Tumor Associated Macrophages, as the Dominant Immune Cells, Are an Indispensable Target for Immunologically Cold Tumor-Glioma Therapy? Front Cell Dev Biol, 2021. 9: p. 706286.

36. Mojsilovic, S.S., et al., The Metabolic Features of Tumor-Associated Macrophages: Opportunities for Immunotherapy? Anal Cell Pathol (Amst), 2021. 2021: p. 5523055.

37. Lopez-Yrigoyen, M., L. Cassetta, and J.W. Pollard, Macrophage targeting in cancer. Ann N Y Acad Sci, 2021. 1499(1): p. 18–41.

38. Wu, Q., et al., Macrophage biology plays a central role during ionizing radiation-elicited tumor response. Biomed J, 2017. 40(4): p. 200–211.

39. Groves, A.M., et al., Effects of IL-4 on pulmonary fibrosis and the accumulation and phenotype of macrophage subpopulations following thoracic irradiation. Int J Radiat Biol, 2016. 92(12): p. 754–765.

40. Jarosz-Biej, M., et al., Tumor Microenvironment as A “Game Changer” in Cancer Radiotherapy. Int J Mol Sci, 2019. 20(13).

41. Zarif, J.C., et al., Mannose Receptor-positive Macrophage Infiltration Correlates with Prostate Cancer Onset and Metastatic Castration-resistant Disease. Eur Urol Oncol, 2019. 2(4): p. 429–436.

42. Giunchi, F., M. Fiorentino, and M. Loda, The Metabolic Landscape of Prostate Cancer. Eur Urol Oncol, 2019. 2(1): p. 28–36.

43. Zhang, J., et al., Meta-Analysis of the Prognostic and Clinical Value of Tumor-Associated Macrophages in Hepatocellular Carcinoma. J Invest Surg, 2019: p. 1–10.

44. Li, J., et al., Prognostic impact of tumor-associated macrophage infiltration in esophageal cancer: a meta-analysis. Future Oncol, 2019. 15(19): p. 2303–2317.

45. Siefert, J.C., et al., The Prognostic Potential of Human Prostate Cancer-Associated Macrophage Subtypes as Revealed by Single-Cell Transcriptomics. Mol Cancer Res, 2021.

46. Pollard, J.M. and R.A. Gatti, Clinical radiation sensitivity with DNA repair disorders: an overview. International journal of radiation oncology, biology, physics, 2009. 74(5): p. 1323–1331.

47. Gold, L., et al., Aptamer-based multiplexed proteomic technology for biomarker discovery. PLoS One, 2010. 5(12): p. e15004.

48. Liu, M., et al., Humoral autoimmune response to nucleophosmin in the immunodiagnosis of hepatocellular carcinoma. Oncol Rep, 2015. 33(5): p. 2245–52.

49. Helwa, I., et al., A Comparative Study of Serum Exosome Isolation Using Differential Ultracentrifugation and Three Commercial Reagents. PLoS One, 2017. 12(1): p. e0170628.

